# Mothers’ experiences of early interaction in multiple pregnancies: A qualitative interview study

**DOI:** 10.64898/2026.09.15.26363150

**Authors:** Petra Holopainen, Päivi Kankkunen, Kristiina Heinonen

## Abstract

**Background:** Forming an equal attachment relationship with more than one child at a time can be challenging for parents. The risks associated with multiple pregnancies for the expecting parent and fetuses can adversely affect the entire pregnancy and influence the early interaction between the woman and the baby. The purpose of this empirical study was to describe from the perspective of women which factors promote and weaken early interaction in multiple pregnancies.

**Methods:** Data collection was conducted remotely through semistructured individual interviews, in collaboration with a regional welfare agency and multiple birth family associations. A total of 11 women expecting twins voluntarily participated in the interviews. The qualitative data were analysed via inductive content analysis.

**Results:** Factors that strengthened early maternal–fetal interaction included considering the fetus and fetuses as part of the family during pregnancy, naming the fetuses individually, and using terms other than “fetus.” Strong social support and parents’ understanding of fetal awareness and reciprocity also significantly promoted early interaction. Conversely, early knowledge of expecting twins, insufficient guidance from healthcare professionals for the formation of early interactions, unequal attachment to fetuses, a lack of knowledge, and fears weakened early interactions.

**Conclusions:** Our findings suggest that early interaction and guidance should be initiated as soon as parents learn that they expect more than one child. Throughout prenatal visits, there should be a focus on supporting the resources of pregnant women and their families to enhance maternal capabilities.

## Background

Twins and triplets account for 2–3% of births worldwide, with the incidence increasing due to fertility treatments and increasing maternal age^1–2^. The prevalence of multiple pregnancies varies substantially across regions and populations, particularly for dizygotic twinning, which is more common in parts of Africa and less frequent in Asia^3^. Globally, approximately 80% of multiple pregnancies occur spontaneously, influenced by factors such as maternal family history, multiparity, and tall stature^4^.

Finland provides an example of a high-performing maternal health system, where universal healthcare, midwife-led maternity clinics, and routine prenatal screening ensure comprehensive follow-up for pregnant women. In multiple pregnancies, ultrasound examinations are performed more frequently, and care is coordinated between primary maternity clinics and specialized hospital units. This system enables early identification of risks such as chorionicity, fetal growth restriction, and preterm labor, contributing to Finland’s consistently strong maternal and neonatal outcomes^8^.

Pregnancy and birth are transitional phases in the life of parents, marking the beginning of their journey into parenthood, where they develop new roles, responsibilities, and identities^5^. Multiple pregnancies are high-risk for both women and fetuses. Ultrasound exams determine whether fetuses share a placenta which impacts how pregnancy is managed and monitored. Compared with dizygotic twins, monozygotic twins who share a placenta face greater risks.^6^ The key risk factors for fetuses include preterm birth, low birth weight^7^ and fetal death^6^. Premature births in Europe and the U.S. range from 50–62%^8,9^.

Early interaction, the bond formed between caregivers and children, differs across multiple pregnancies. It may take longer to form an attachment to each fetus.^10^ The womb provides a unique sensory environment, helping fetuses recognize familiar voices^11^. Mothers bond with multiple children as individuals and as twins, often forming mental images once they learn that they are expecting twins^12^. Identical twins may be viewed collectively, complicating individual attachment^13^.

Learning about multiple pregnancies can be shocking, and related risks can delay bonding. Anxiety about fetal health or sharing a placenta can hinder attachment.^14^ Factors such as advanced maternal age, depression, low self-esteem, relationship difficulties, and demands from older children further impact early interaction^15–18^.

While most research on early interaction in multiple births focuses on the postpartum period^12,19^, studying this during pregnancy is essential. This study provides insights into how mothers experience and develop relationships with their fetuses and may help identify ways to support families during multiple pregnancies and promote family well-being after childbirth^20^. Therefore, the purpose of this study was to describe mothers’ perspectives on early interactions with their fetuses during multiple pregnancies and to provide knowledge about the characteristics of these interactions.

## Methods

### Study design and participants

The aim of this study was to describe mothers’ perspectives on early interactions with their fetuses during twin pregnancies. This qualitative descriptive study was conducted in Finland as part of the TWIN LIFE 2021–2026 research project, Nurses’ Competence in Multiple-Birth Family Nursing.

The data were collected through individual semistructured thematic interviews. The participants were women expecting twins at various stages of pregnancy. A total of 11 women participated voluntarily in the study and were interviewed remotely. In the reporting of this study, COREQ guidelines were utilized to increase the transparency and replicability of the qualitative research. The study was reported in accordance with the Consolidated Criteria for Reporting Qualitative Research (COREQ) guidelines to enhance the transparency of the research process.^21^

### Researcher role and reflexivity

The research team maintained reflexivity throughout the study. The interviewer, a midwife and maternal health researcher, acknowledged that her professional background could influence data collection and interpretation. To minimize this, the team discussed pre-understandings and compared interpretations to ensure that themes reflected participants’ experiences rather than researcher assumptions.

### Data collection

Participants were recruited in collaboration with maternity clinics and Finnish multiple-birth associations between November 2023 and February 2024. Nurses distributed study information during appointments. The local association also emailed the information to their members. A social media campaign was launched in December 2023 to further promote participation.

All interested participants contacted the researcher by themselves and received more information about the study before providing their informed consent. They could withdraw from the study at any time without needing to give a reason. The interviews were conducted using Teams and were recorded with a recorder and Teams’ recording feature, which lasted between 28 minutes and 1 hour 12 minutes.

### Data analysis

The data were transcribed verbatim and analysed via inductive content analysis. The transcribed text totaled 167 pages. The data were pseudonymized by removing direct identifiers. In the analysis process, expressions relevant to the research questions were identified from the data, grouped into similar categories and finally abstracted into main categories. This analysis method was chosen because of the limited prior research on early interactions in twin pregnancies. Throughout the process, original statements were numerically coded for traceability, and findings were iteratively reviewed to ensure accuracy and reliability^22^. The analysis led to the identification of five main categories, with a specific focus on factors that promote and hinder early interaction. The results of this analysis are reported in detail via descriptive text, tables, and figures to convey the findings comprehensively. An example of the progression of the analysis can be seen in Table 1.

**Table 1.** An example of inductive analysis from simplified expression into the main category. The unifying category is not presented.

| Simplified expression | Subcategory | Main category |
| --- | --- | --- |
| • Concern about high-risk pregnancy<br>• Concerns<br>• Specific features related to multiple pregnancies are concerning<br>• Concern is the biggest issue | Concerns about multiple pregnancy | Challenges posed by multiple pregnancies |
| • Afraid to prepare for birth<br>• Concern about the development of fetuses<br>• Knowledge of Mo-Di pregnancy increased concern<br>• Worry, if one does not develop | Concerns about the well-being of the fetuses |  |
| • Fear of high-risk pregnancy<br>• Knowledge increases distress<br>• Anxiety and fear about check-ups | Fears caused by multiple pregnancy |  |
| • Fear of premature birth<br>• Concern about premature birth | Fear of prematurity |  |
| • Fear if something happens to one<br>• Fear of miscarriage<br>• Excluding the pregnancy from mind before the nuchal translucency ultrasound<br>• Desire to hold back attachment<br>• Fear of loss<br>• Fear of losing one or both fetuses<br>• Concern if one is in distress | Fear of losing the fetus or fetuses |  |

## Results

### Sample characteristics

The participants were women aged 28 - 39 years (mean age 34). At the time of the interviews, the gestational age ranged from 17+6 to 36+4 weeks. All the participants were in a relationship; two were cohabiting and nine were married. Five participants were expecting twins ad their first children. Educational backgrounds included secondary education as well as lower and higher university degrees. Most of the participants had completed a higher university degree.

### Findings

Three main themes emerged as factors facilitating early interaction: (1) the sensory environment in the formation of early interaction, (2) reciprocity in early interaction, and (3) preparation for interaction between the mother and the fetuses (see Figure 1). Two main themes emerged as factors hindering early interaction: (1) maternal challenges in forming early interactions in multiple pregnancies, and (2) challenges posed by multiple pregnancies to the formation of early interactions (see Figure 2). Overall, these findings indicate that early maternal–fetal interaction in multiple pregnancies are shaped by both facilitating and hindering factors. The relationships between these factors and mothers’ experiences are summarized in Figure 3.

**Figure 1.**
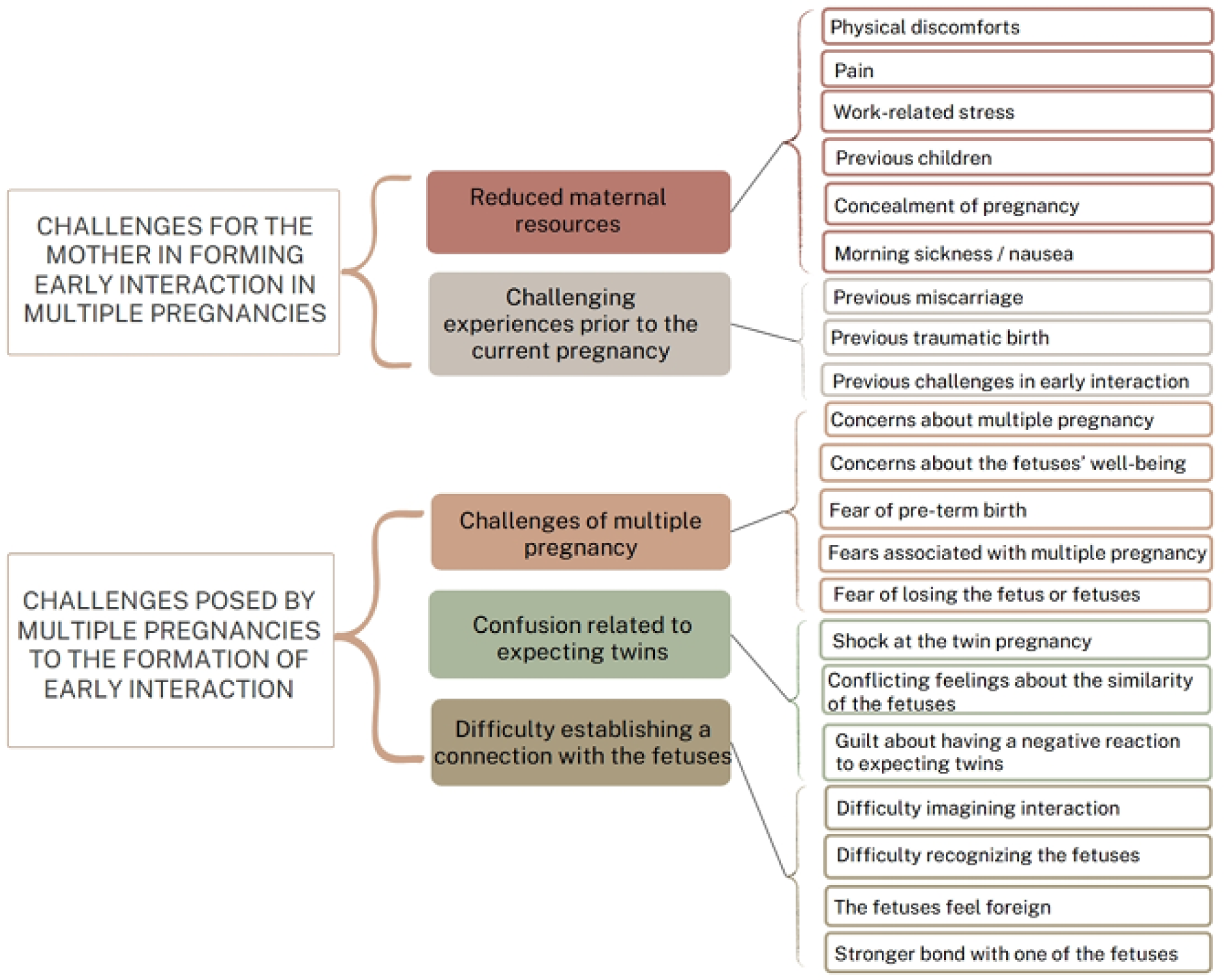
Factors hindering early interaction

**Figure 2.**
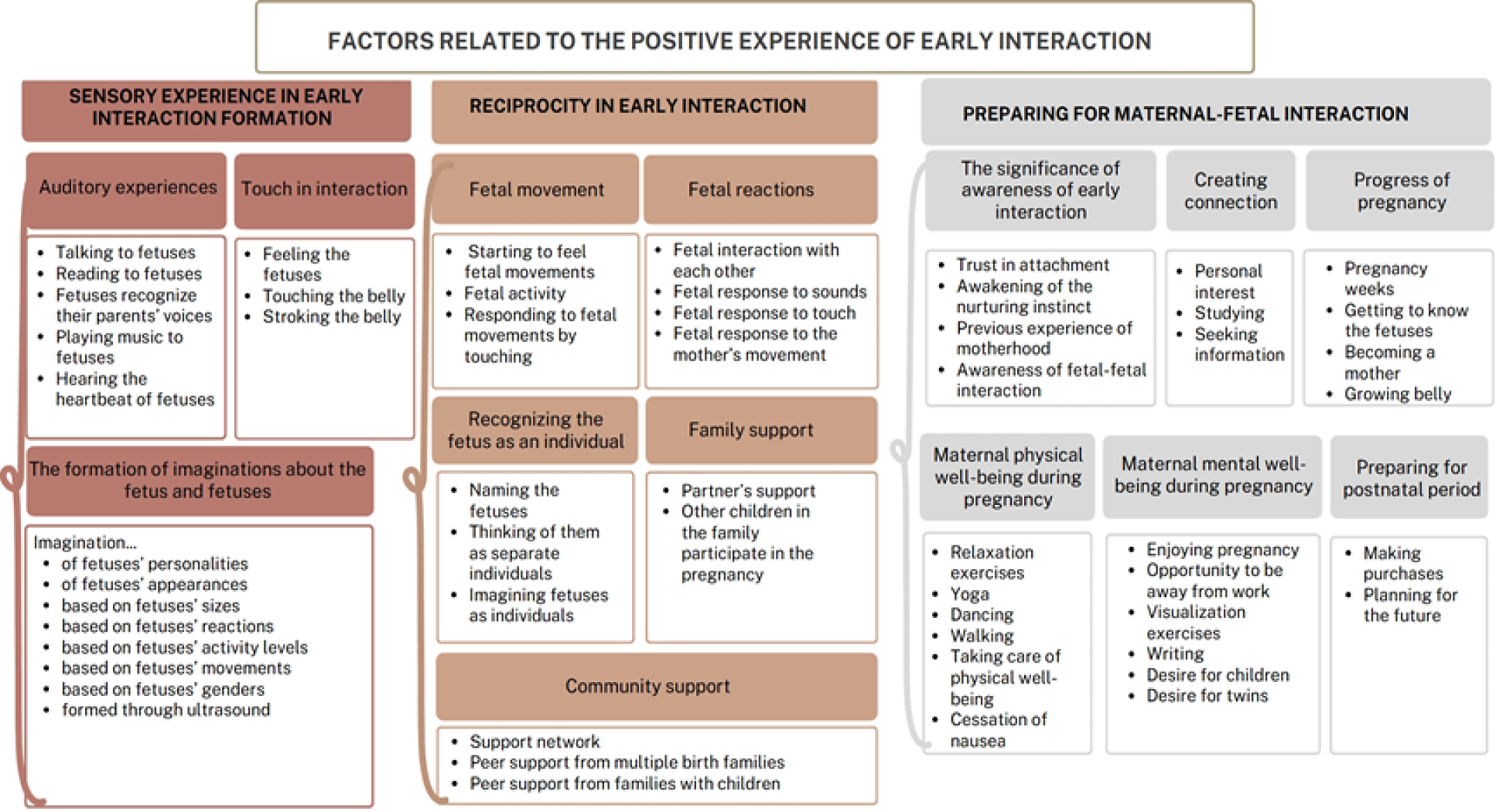
Factors facilitating early interaction

**Figure 3.**
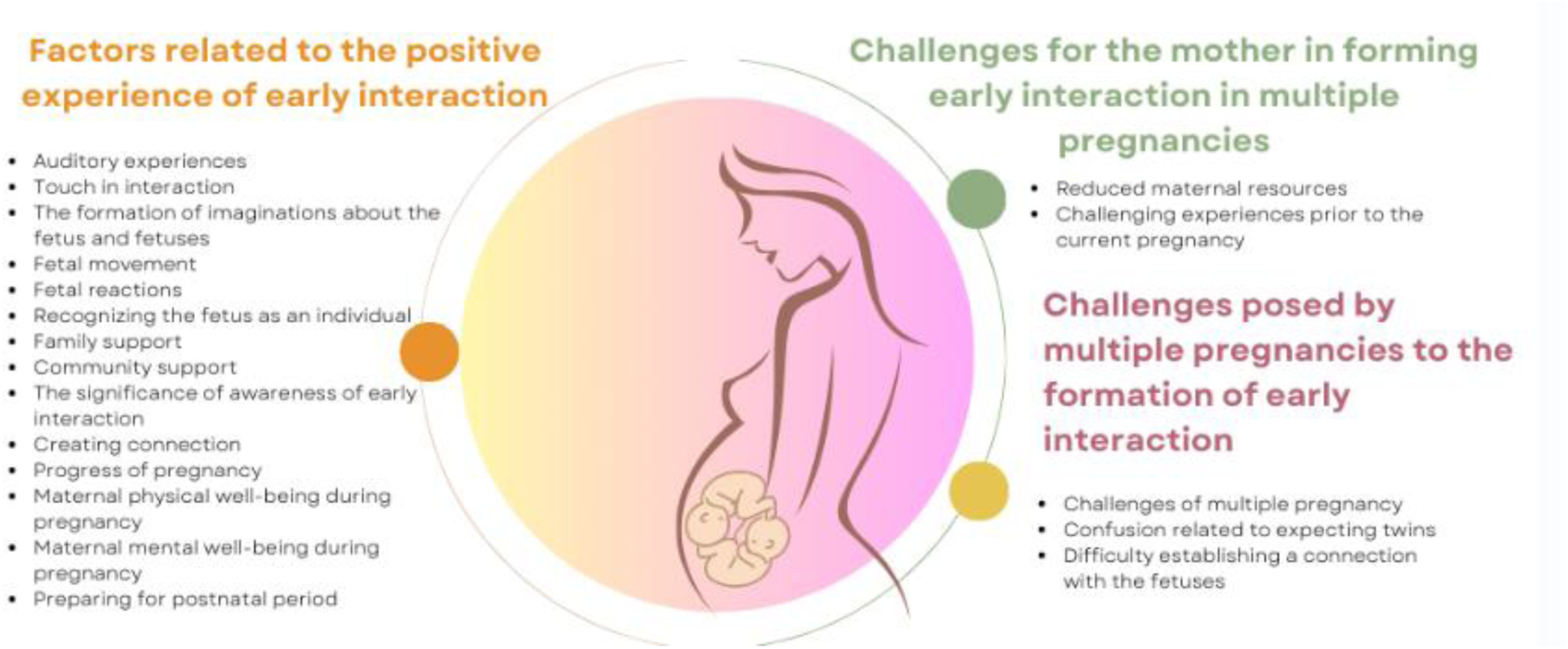
Mothers’ experiences of early interaction in multiple pregnancy (author-created figure).

### Sensory experience in early interaction formation

Mothers interact with their fetuses through various sensory modalities. Speaking and reading to their fetuses are believed to create a calming intrauterine environment. Mothers also played music and moved to its rhythm, which they believed would help fetuses recognize music later.

> *“We relax together, listening to some music -- I’ve tried playing the same songs so that they would recognize them.” (2)*

Fetal heartbeats are important to mothers, as they deepen their sense of having real babies. “*-- then there are prenatal visits, and -- hearing the heartbeats.” (7)* Additionally, mothers noted that the awareness that the fetuses could hear their voices helped them direct their speech toward the fetuses specifically, rather than just the abdomen.

Touching was another way mothers formed connections with their fetuses. They described touching as a form —stroking, massaging, and feeling for movements— to create bonds. This physical contact focused on the abdomen and occurred throughout the pregnancy. In the second and third trimesters, mothers hoped for a response from the fetuses as movement and kicking increased.

Nearly all mothers highlighted that creating mental images of fetuses was one of the most significant ways to promote early interaction. Mothers establish a foundation for interaction by conceptualizing fetuses as babies and discussing these images with their partners. Conscious relaxation and visualization enhanced the bonding process with one or both fetuses simultaneously. Mothers form images on the basis of the personalities, appearances, and sizes of fetuses, often envisioning them as happy, playful, cute, and small. Differences in fetal size led to distinct mental images of the fetuses as individuals.

> *“We have like talked with my husband about the babies and wondered what they’re like.” (1) “A was 200 grams smaller than B -- which is a significant difference -- perhaps 11%. This creates different images of them as individuals.” (6)*

Mothers reported that knowing the fetuses’ sex and having ultrasound examinations significantly enhanced early interaction. This knowledge helps them envision the personalities of fetuses and their own journey toward motherhood. Ultrasound images reinforced the reality of carrying two babies.

> *“-- early bonding -- significantly easier -- it is so important to see them on the screen. In addition, that everything is fine there.” (8)*

### Reciprocity in early interaction

Mothers identified fetal movements, typically felt before the 20th week of pregnancy, as a key turning point. These movements marked the beginning of significant emotional connection. Responding to movements through touch further enhanced this interaction, leading to increased attachment when mothers perceived a reciprocal response. Mothers focus on developing individual relationships with each fetus, often naming them to recognize them as separate individuals. As the pregnancy progressed, their understanding of each fetus strengthened, and they looked forward to seeing if the imagined characteristics matched reality.

> *“I always want to touch, so if they react somehow, I want to tell them that I’m here -- it helps me feel emotionally present,” (2)*
>
> *“-- then, when I started feeling the kicks, it also made it more concrete -- that there are actually - - not just one but two babies there, leading to more feelings and attachment to them.” (6)*
>
> *“Initially, we used a shared name for the twins when talking about them. Around the midpoint of the pregnancy -- we were encouraged to name them separately.” (2)*

Social networks play a crucial role. Partner support, including attending prenatal visits and engaging with fetuses, positively impacts the bonding process. Involving other children in the pregnancy was also significant; older siblings participated by feeling movements and discussing names.

> *“Positive humor from older children provided extra strength to cope with pregnancy” (10)*
>
> *“What has facilitated early interaction is also that my husband has been very -- present, even coming to appointments” (2)*

### *Preparing for* maternal‒fetal interaction

Mothers identified key facilitators of early interaction: awareness, conscious connection, pregnancy progression, maternal well-being, and postbirth preparation. They viewed early interaction as a natural aspect of motherhood that develops easily with focus and without pressure. Trust in maternal instincts and previous experiences facilitated this, while understanding the connection between the fetuses in the womb reinforced attachment. Mothers sought information on twin pregnancies through articles and family association websites Pregnancy progression influences early interactions. In the first trimester, mothers experienced bonding fears, but connections strengthened in the second trimester. As pregnancies advanced and their abdomen grew, mothers became more prepared, making the experience feel more concrete.

> *“Now in the second trimester, it feels like the further the pregnancy progresses, the more concrete the babies become” (4)*
>
> *“-- the information from the twin family association -- all the facts available -- clearly increased the attachment and anticipation for the future.” (8)*
>
> *“We’ll manage, and -- the attachment will form with both.” (1)*

Physical and psychological well-being are crucial. Mothers practiced relaxation, yoga, dancing, and walking to increase their well-being and connection with fetuses. The cessation of morning sickness marked a positive milestone. Enjoying the pregnancy, engaging in visualization, writing, and leaving work prior to maternity leave reduced stress allowing for greater focus on the pregnancy and fetuses.

> *“Then I do yoga and I feel like it strengthens the connection between us.” (11)*

### Challenges for the mother in forming early interactions in multiple pregnancies

Mothers face diminished resources because of physical discomfort and pain, making it difficult to focus on pregnancy. Stress from work, demands from older children, and past traumatic experiences, such as miscarriages, heightened fears and complicated early bonding. (Fig. 2)

> *“I had very strong nausea in the early stages of pregnancy, and I think that truly affected me, because when you feel unwell, you don’t truly want to -- even think about it.” (8)*
>
> *“We have one miscarriage in the past, so I was afraid that the other one might miscarry or that both might.” (2)*

### Challenges posed by multiple pregnancies to the formation of early interactions

Mothers experienced heightened anxiety regarding the risks of multiple pregnancies, fear of complications, preterm birth, or the loss of one or both fetuses. This anxiety delayed preparations and hindered early bonding. To protect themselves emotionally, mothers used medical terms such as “embryos” and “fetuses” rather than “babies” until later in the pregnancy.

The initial discovery of expecting twins caused shock, distress, and guilt, although these feelings typically became more positive over time. Mothers expecting identical twins expressed mixed emotions, worrying about the children’s individuality. Early interaction was challenged by the unfamiliarity of the fetuses, difficulty in forming mental images, and struggles with unequal attachment or difficulty in distinguishing between the twins.

> *“At first, we only talked about -- embryos and fetuses and not about babies at all -- I definitely used those terms consciously, just to kind of hold back a bit -- not daring to think too much about the future yet.” (8)*

## Discussion

This study examined the factors influencing early maternal‒fetal interactions in multiple pregnancies, providing insights into the unique experiences of mothers expecting twins. A central finding was that fetal movements, typically felt before the 20th gestational week, marked a significant turning point in early bonding. This finding underscores the importance of gestational progression in the development of attachment, which aligns with previous research that demonstrated a strong correlation between gestational age and maternal‒fetal interaction^15,19,23–24^. However, mothers of multiples face distinct challenges, including a higher risk of preterm birth and consequently less time for psychological adaptation to parenthood^8^. Therefore, timely support from healthcare professionals is crucial to foster bonding throughout pregnancy.

Prenatal monitoring, including ultrasounds and fetal heartbeat checks, was identified as a significant facilitator of early interaction. Visualizing and hearing fetuses strengthens maternal mental representations, supporting the development of individualized relationships with each fetus^23,25^. Furthermore, knowledge of the fetuses’ biological sex enhances bonding by allowing mothers to create personalized narratives and differentiate between the children, which is consistent with prior findings^23,26^. Nevertheless, since the impact of sex disclosure on bonding can vary^15,18^, it remains essential for prenatal care to respect parental preferences.

Social support emerged as a cornerstone of early interaction. Support from partners, older children, and wider networks provided emotional stability and opportunities for reflection. Such support allows mothers to focus on bonding while reducing stress, which aligns with the literature regarding the role of social networks in fostering secure attachment^27^. Notably, this study did not include single parents; exploring their unique challenges in establishing early interaction remains a vital area for future research.

Mothers utilize various sensory modalities —such as touch, speech, music, and visualization—to enhance bonding. Activities such as repetitive reading or listening to music enable mothers to feel reciprocal engagement, supporting attachment to both children simultaneously^25–26,28^. These strategies are not yet systematically integrated into prenatal guidance for families expecting multiples, yet they would be an important addition as tools to help parents establish early interactions during pregnancy.

Despite these facilitators, challenges unique to multiple pregnancies were evident. Maternal anxiety regarding fetal risk, physical discomfort (such as severe nausea), and the initial emotional shock of a twin diagnosis were found to hinder attachment. These findings are consistent with previous research indicating that heightened maternal stress and physical symptoms in multiple gestations can complicate the early bonding process^29^. Furthermore, unequal attachment to one fetus—often owing to perceived differences in activity—illustrates the potential influence of “monotropy,” where a parent initially bonds more strongly with one child^10,30^. This aligns with the established literature on maternal preferential attachment and the complexities of establishing balanced relationships with multiple fetuses^15,26,31–32^.

The generalizability of these findings must be considered in light of the Finnish context, where participants had access to universal healthcare, regular prenatal monitoring, and high-quality maternity services. These structural conditions likely facilitated early interaction by reducing uncertainty, enabling frequent contact with healthcare professionals, and supporting maternal well-being. In many low- and middle-income countries, where access to ultrasound examinations, continuity of care, and psychosocial support is limited, mothers may face greater barriers to forming early maternal–fetal relationships.^7^ Therefore, while the findings offer important insights into early interaction in multiple pregnancies, they may not fully reflect the experiences of mothers in settings with fewer resources or higher perinatal risks. Future research should explore how early interaction develops in diverse global contexts to better understand how health system structures shape bonding in multiple pregnancies.

Finally, the quality and continuity of prenatal care significantly influence the bonding process. Mothers valued familiar healthcare providers and clear explanations during monitoring, which increased participation and confidence. Conversely, visits focused strictly on medical measurements left mothers feeling excluded. These results highlight the need for individualized, relationship-centered care and specialized professional knowledge in supporting multiple-birth families.

## Conclusion

This study explored and described factors influencing early maternal–fetal interaction during multiple pregnancies from mothers’ perspectives. The findings indicate that early maternal–fetal interaction is a complex and dynamic process that occurs during pregnancy and is influenced by both maternal experiences and the perceived presence and characteristics of the fetuses. Fetal movements, prenatal monitoring, knowledge of fetal sex, social support, and sensory engagement appeared to facilitate interaction, whereas maternal anxiety, physical discomfort, and the psychological demands associated with a high-risk pregnancy could hinder it. These findings contribute to the understanding of early maternal–fetal interaction during multiple pregnancies, an area that has received less attention than the postpartum period.^12,19^

The findings highlight the important role of healthcare professionals in supporting mothers and families expecting multiples. Timely and individualized guidance may help mothers recognize and respond to opportunities for interaction with each fetus and support the development of an individual relationship with each child. Such support should take into account the unique characteristics and challenges associated with multiple pregnancies and should extend to the family as a whole. This is particularly relevant given the increased psychological and physical demands associated with multiple pregnancies.14–18 These findings may inform prenatal care, health education, and family-centred support for families expecting twins or other multiples.

Further research is needed to examine early maternal–fetal interaction in different family contexts, including single-parent families, and to evaluate interventions designed to support interaction with each fetus. Longitudinal research could further clarify how maternal–fetal interaction develop throughout pregnancy and how they are related to postnatal bonding and parent–infant relationships after birth. Such research could contribute to the development of evidence-based approaches to prenatal care and support for families expecting multiples.

## Data Availability

The datasets generated and analysed during the current study are not publicly available owing to the sensitive nature of the interview data and restrictions related to participant confidentiality and data protection.

## List of abbreviations

COREQ: Consolidated Criteria for Reporting Qualitative Research

## Declarations

### Ethics approval and consent to participate

This study received ethical approval from the University of Eastern Finland Research Ethics Committee on 22 May 2023 (Statement 24/2023). The study was conducted in accordance with the ethical principles of the 1964 Declaration of Helsinki^33^. All participants received information about the study and provided informed consent to participate. Participation was voluntary, and participants were informed of their right to withdraw from the study at any time without providing a reason. Participant confidentiality and anonymity were maintained throughout the research process.

To enhance the rigor of the study, a pilot interview was conducted to identify any necessary adjustments to the interview guide^34^. The primary supervisor observed the pilot interview, and as no significant changes were needed, the pilot interview data were included in the final dataset. The research process and data analysis were discussed regularly with the supervisory team to support methodological rigor and reflexivity throughout the study.

### Consent for publication

Not applicable

### Authors’ contributions

**PH** contributed to the conceptualization of the study, data collection, and inductive content analysis. **KH** and **PK** contributed to the conceptualization of the study and supervised the research process. **PH** drafted the manuscript, and **PH, KH, and PK** contributed to reviewing and editing the manuscript. All the authors have read and approved the final manuscript.

## Acknowledgements

The authors would like to thank the Finnish Multiple Births Association and a Finnish wellbeing services county for their collaboration in this study. We would also like to thank all the women expecting twins who voluntarily participated in the study and who generously shared their experiences.

